# The association between treatment with heparin and survival in patients with Covid-19

**DOI:** 10.1101/2020.05.27.20114694

**Authors:** Luis Ayerbe, Carlos Risco, Salma Ayis

## Abstract

**Objectives:** This study investigates the association between the treatment with heparin and mortality in patients admitted with Covid-19.

**Methods:** Routinely recorded, clinical data, up to the 24^th^ of April 2020, from the 2075 patients with Covid-19, admitted in 17 hospitals in Spain between the 1st of March and the 20th of April 2020 were used. The following variables were extracted for this study: age, gender, temperature, and saturation of oxygen on admission, treatment with heparin, hydroxychloroquine, azithromycin, steroids, tocilizumab, a combination of lopinavir with ritonavir, and oseltamivir, together with data on mortality. Multivariable logistic regression models were used to investigate the associations.

**Results:** At the time of collecting the data, 301 patients had died, 1447 had been discharged home from the hospitals, 201 were still admitted, and 126 had been transferred to hospitals not included in the study. Median follow up time was 8 (IQR:5-12) days. Heparin had been used in 1734 patients. Heparin was associated with lower mortality when the model was adjusted for age and gender, with OR (95%CI): 0.55 (0.37-0.82) p=0.003. This association remained significant when saturation of oxygen <90%, and temperature >37C were added to de model with OR: 0.54(0.36-0.82) p=0.003, and also when all the other drugs were included as covariates OR: 0.42 (0.26-0.66) p<0.001.

**Conclusions:** The association between heparin and lower mortality observed in this study can be acknowledged by clinicians in hospitals and in the community. Randomized controlled trials to assess the causal effects of heparin in different therapeutic regimes are required.

- The administration of heparin was associated with lower mortality in patients admitted with Covid-19.
- Our findings support that there is a thrombotic component in the development of respiratory distress for these patients.
- The positive effect of heparin seems consistent and its use, when indicated, could be considered in clinical settings.
- Randomized controlled trials are necessary to complement observational studies, and assess the causal associations between heparin, in different therapeutic regimes, and clinical outcomes.
- Heparin is easy to administer, its use in ambulatory patients, to prevent admissions, or reduce their duration, could also be considered by clinicians and future researchers.

## Introduction

In December 2019 an outbreak of disease caused by the virus SARS-CoV-2 was declared in China. This disease, later named Covid-19, spread throughout the world in the first few months of 2020.[1, 2] Around 80% of patients have mild symptoms, the rest can develop severe disease, mainly interstitial pneumonia and acute respiratory distress syndrome, and require hospital admission. Intensive care may be necessary for 5% of patients. Mortality is estimated to be less than 2%. [3, 4] The large number of simultaneous cases of Covid-19 has overloaded hospitals, making it very difficult to provide an adequate care. The spread of Covid-19 is negatively affecting all areas of healthcare, and is also having an adverse impact on the entire society and the international economy.

Thrombotic events in different tissues, with consistent clinical laboratory and radiological findings, have been reported in patients with Covid-19.[5-7] Perfusion abnormalities in lungs have been observed in patients who underwent dual-energy CTs.[8] There is also pathological evidence of the presence of platelet-fibrin thrombi in small arterial vessels in lung tissues of patients who have died of Covid-19.[9] Based on these finding, doctors have used heparin to treat or prevent the coagulation disorder associated with the infection.[5-7, 9] This treatment can be associated with the clinical improvement of the patients, and decrease the duration of admissions and the mortality of Covid-19. Heparin is safe, economical, and easy to use, it can be given both to admitted and outpatients. However, the use of heparin in Covid-19 patients is still supported by limited evidence. This study investigates the association of the use of heparin with mortality in a large number of patients admitted with Covid-19.

## Methods

Anonymized clinical records up to the 24^th^ of April 2020, of all patients (n=2075) with Covid-19, admitted in 17 Spanish hospitals, of the healthcare provider HM Hospitales, were reviewed. These hospitals are based in the provinces of Barcelona, Coruña, León, Madrid, Pontevedra, and Toledo.[10] Patients had been diagnosed with polymerase chain reaction test of respiratory samples for SARS-CoV-2, between the 1st of March and the 20th of April 2020. Seven patients had been admitted before the 1st of March 2020. Data on age, gender, temperature and saturation of oxygen at the time of admission, together with treatments at any time during admission with heparin, hydroxychloroquine, azithromycin, steroids, tocilizumab, a combination of lopinavir with ritonavir, and oseltamivir, were collected. The limited evidence on which these treatments were based, and the rapidly evolving clinical protocols, resulted in these drugs being given in many different specific preparations, doses, and frequency. No information on dosage, duration of treatment, route of administration, or specific preparations of these drugs was collected. Finally, data on mortality were recorded.

The age of patients treated and not treated with heparin was compared with t tests. The proportion of men and women for those treated and not treated with heparin was compared with chi squared tests. The association between the treatment with heparin and mortality was examined with a logistic regression model that was first adjusted for age and gender. Second, two markers of severity, temperature > 37 C and saturation of oxygen < 90% on admission, were introduced into the model as well. Finally, treatments with hydroxychloroquine, azithromycin, steroids, tocilizumab, a combination of lopinavir with ritonavir, or oseltamivir, were added to the model. Sensitivity analyses were made to compare estimates based on using missing data of severity markers as categories with, 1) estimates based on complete data dropping variables with missing observations, and 2) complete case analysis. Stata 14 was used for the analysis.[11] The ethics committee of HM Hospitales approved this study. Data were anonymized before the authors could access it.

## Results

Among the 2075 patients whose records were reviewed, 1256 were men, 819 were women, and the mean age was 67.57. At the time of collecting the data, 301 patients had died, 1447 had been discharged home from the hospitals, 201 were still admitted, and 126 had been transferred to hospitals not included in the study. Median follow up time was 8 (IQR: 5-12) days. Among the 1734 patients who received heparin 242 had died. The patients treated with heparin were older than the ones who did not received it (p<0.001) and the proportion of men and women who received heparin did not have significant difference. (Table 1).

**Table 1.** Description of the patients included in the study

|  |  | N | Age M(SD) | Female<br>n(%) | Death n(%) |
| --- | --- | --- | --- | --- | --- |
| Total cohort |  | 2075 | 67.57(15.52) | 819<br>(39.47%) | 301 (14.51%) |
| Oxygen Saturation<90% | Yes | 70 | 73.18(13.7) | 20 (28.57) | 28(53.85) |
|  | No | 221 | 67.00(16.14) | 95 (42.99) | 24(46.15) |
| Temperature > 37 C | Yes | 159 | 61.20(17.27) | 47(29.56) | 24 (10.86) |
|  | No | 1422 | 68.53(15.65) | 577(40.58) | 197 (89.14) |
| Heparin | Yes | 1734 | 68.77(15.09) | 686(39.56) | 242(13.96) |
|  | No | 285 | 61.76(17.67) | 96(33.68) | 44(15.44) |
| Hydroxychloroquine | Yes | 1857 | 67.11(15.51) | 705(37.96) | 237(12.76) |
|  | No | 162 | 73.47(16.22) | 77(47.53) | 49(30.25) |
| Azithromycin | Yes | 1223 | 68.33(15.03) | 456(37.29) | 146(11.94) |
|  | No | 796 | 66.54(16.51) | 326(40.95) | 140(17.59) |
| Steroids | Yes | 960 | 69.88(16.76) | 330(34.38) | 200(20.83) |
|  | No | 1059 | 65.58(16.76) | 452(42.68) | 86(8.12) |
| Tocilizumab | Yes | 421 | 66.1(13.11) | 117(27.79) | 89(21.14) |
|  | No | 1598 | 68.00(16.24) | 665(41.61) | 197(12.33) |
| Lopinavir+Ritonavir | Yes | 1230 | 63.94(14.28) | 421(34.23) | 160(13.01) |
|  | No | 789 | 73.37(15.99) | 361(45.75) | 126(15.97) |
| Oseltamivir | Yes | 132 | 67.78(13.79) | 51(38.64) | 26(19.70) |
|  | No | 1887 | 67.61(15.78) | 731(38.74) | 260(13.78) |

Heparin was associated with lower mortality when the model was adjusted for age and gender, with OR (95%CI): 0.55 (0.37-0.82) p=0.003. This association remain significant when saturation of oxygen <90%, and temperature >37C were added to de model with OR: 0.54(0.36-0.82) p=0.003, and also when all the other drugs were included as covariates OR: 0.42 (0.26-0.66) p<0.001.

## Discussion

The administration of heparin was associated with lower mortality in patients admitted with Covid-19. This study has strengths and limitations. Patients were not randomized and the difference in outcome may be explained by factors other than the treatment with heparin. In an effort to control for the severity of disease, or the effects of other drugs, the models were adjusted for clinical markers and other treatments. The observation of a large number of unselected patients in 17 hospitals, and the analyses run with some variations to test the consistency of the results, are also strengths of this study. However, residual confounding is always present in all observational research. For all models the sensitivity analyses described in the methods were used to assess the impact of missing data on the two severity markers, and results were consistent with those presented.

While the nature of the hypoperfusion of lungs in Covid-19 patients is still not fully understood, our findings support that there is a thrombotic component in the development of respiratory distress for these patients.[6, 8] The association between heparin and survival is consistent with the findings of an observational study where among the 99 patients who received heparin, and had a sepsis-induced coagulopathy index>4, mortality was significantly lower.[12] A recent observational study conducted in a hospital has also reported lower mortality among the 786 Covid-19 patients who received anticoagulation.[13]

To the best of our knowledge, there is no interventional evidence on the management of the coagulopathy associated with Covid-19. The positive effect of heparin seems consistent and its use, when indicated, could be considered in clinical settings. Randomized controlled trials are necessary to complement observational studies, and assess the causal associations between heparin, in different therapeutic regimes, and clinical outcomes. Heparin is easy to administer, its use in ambulatory patients, to prevent admissions, or reduce their duration, could also be considered by clinicians and future researchers.

## Data Availability

Queries on data should be addressed to HM Hospitales

## Declarations

### Ethics approval

The Ethics committee of HM Hospitales approved this study.

### Consent to participate

All data were anonymized before researchers could access it.

### Consent for publication

Authors have agreed for the final version of this paper to be submitted for publication

### Availability of data and material

Data queries should be addressed to HM Hospitales Authors’ contributions: LA, SA and CR conceived the idea. LA extracted the data. SA conducted the analyses. LA wrote the first draft, that received later contribution from SA and CR.

## Acknowledgement

Thank you to all clinicians and administrators of Hospitales HM (Spain) on which data this study is based.

